# Delay-robust Estimation of the Reproduction Number and Comparative Evaluation on Generated Synthetic Data

**DOI:** 10.1101/2020.11.27.20238618

**Authors:** Benedikt Heidrich, Tillmann Mühlpfordt, Veit Hagenmeyer, Ralf Mikut

## Abstract

The reproduction number is an indicator of the evolution of an epidemic. Consequently, accurate estimators for this number are essential for decision making in politics. Many estimators use filtered data as input to compensate for fluctuations of reported cases. However, for daily-based estimations, this filtering leads to delays. Some approaches use small window sizes for filtering to overcome this issue. This, in turn, leads to an increased periodic behavior of estimators. To overcome these issues, in the present paper, we introduce an estimator for the reproduction number that uses an acausally filtered number of cases as input, hence avoiding both the periodic behavior and the delay. For the filter size, we suggest using a multiple of one week since reported cases often exhibit a weekly pattern. We show that this approach is more robust against periodicities, and that it does not exhibit any delays in the estimation compared to estimators with smaller filter sizes.

Moreover, often it is hard to examine estimators in detail because a ground truth is missing. For analyzing different properties of the estimators, we propose a method to generate synthetic datasets that can be taken as ground truths. Importantly, the synthetic data contains all relevant real-world behavior.

Finally, we apply the proposed estimator to the publicly available coronavirus disease 2019 (COVID-19) data for Germany. We compare the proposed estimator to two estimators used by the federal German Robert Koch Institut (RKI). We observe that our estimator is more stable than the benchmarks especially if the reproduction number is close to 1. Based on the observation that the proposed estimator appears more robust, it may be a useful asset when considering governmental interventions.

The accompanying source code is published under the Apache-2.0 license (https://github.com/timueh/C0VID-19).

## 1 Introduction

Epidemiologists employ the (basic) reproduction number *R*(*t*) at time *t* to quantify the “average number of people infected by a typical case” [9]. Albeit an average number, its importance for policymakers is unquestionable, providing a threshold for the spread of diseases (*R*(*t*) > 1) or their confinement (*R*(*t*) < 1). For instance, many governmental interventions during the coronavirus disease 2019 (COVID-19) pandemic were and are unfortunately based on too-large estimates of the reproduction number *R*(*t*) [8]. Clearly, accurate daily estimates of *R*(*t*) are of paramount relevance for taking further future actions. However, it is also the past that deserves attention: did the estimates of the reproduction number *R*(*t*) justify and show effectiveness of past interventions? Although the existing literature is rich on both kinds of problems, various challenges remain:

- Real-world data is (often) *incomplete*; often the available data pertains only to confirmed cases and to confirmed deaths.
- Moreover, the records are delayed. Figure 1 shows the connection and delays between a model of some disease and the necessary testing and reporting—all of which contributes to the (lagging) reported records.
- A further challenge is that the estimated number of cases 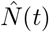 at time *t* might be changed retrospectively. The cause of this effect is the delay in reporting. This delay leads to a changing data basis between two calculations of the nowcasting^1^ model on two subsequent days. Consequently, the result of the nowcasting model changes. Especially for daily-based estimation, this is hard to handle. Thus, in the literature, delay patterns and probabilities of misreporting are used for overcoming this issue. However, such values change during an epidemic. Moreover, since delays are often caused by weekends or public holidays, they lead to periodicities in the reported data, which challenge estimators too.
- Finally, the reproduction number is not observable; it is an artificial, computed number. Consequently, no ground truth is available. However, ground truths are necessary for calculating error measures, and for verifying and justifying a posteriori the decisions that were taken for example by governments. The only observable data are the reported (*C*(*t*)), nowcasted 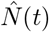 and the estimated number of deaths (compare Figure 1).

**Figure 1:**
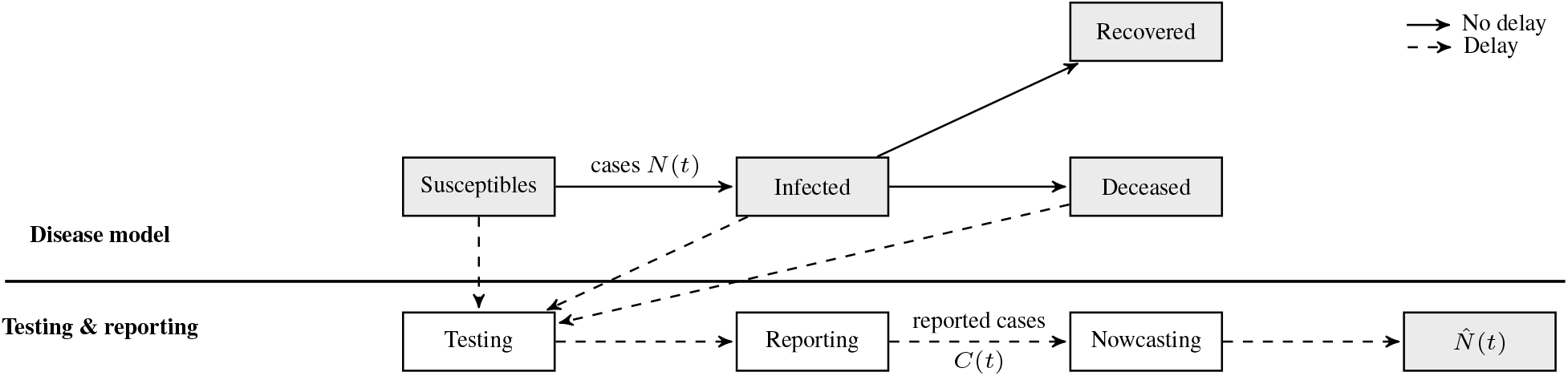
Flow diagram visualizing the roots of delays as well as connections between actual cases, reported cases, and nowcasted cases.

In light of the above challenges, the present paper contributes twofold:

- We provide a periodicity-aware estimator for the reproduction number *R*(*t*). We show that it outperforms two existing approaches in use by the German Robert Koch Institut (RKI) for the raging COVID-19 pandemic. Our proposed estimator uses an acausal filter; it relies on values from the past, the present, and the future. The future values for calculating *R*(*t*) are predicted based on the previous week. This projected acausal filtering can cope with periodicities, much better so than causally filtered approaches with filter sizes less than one week. Also, around the critical threshold of *R*(*t*) ≈ 1 the proposed estimator is more robust.
- We provide a synthetic yet realistic data set, which is based on predefined values of *R*(*t*). Consequently, this data set provides a defined ground truth that allows to compare existing estimators to the proposed one. Moreover, for making the data set more realistic, we incorporate delays that are derived from real-world data about the COVID-19 outbreak in Germany.

The remainder of the paper is structured as follows. Section 2 summarizes related work on *R*(*t*)-estimators. Section 3 introduces the proposed estimator. Subsequently, Section 4 presents the data generation. Afterwards, Section 5 evaluates the estimators on the synthetic dataset. In Section 6, we apply the estimator to the records of the COVID-19 epidemic in Germany. Finally, we wrap up our contributions and give an outlook on future research in Section 7.

## 2 Related Work

There exists a rich literature on estimating the reproduction number *R*(*t*); we merely provide snapshots for the sake of brevity. For instance, Fraser shows how to compute the reproduction number from a combination of historical data and a probability of infection from a case *i* days ago [9]. In contrast, Cintrón-Arias et al. derive the reproduction number *R*(*t*) from the modeling parameters of a standard susceptible-infected-recovered (SIR) model [6]. Instead, Thompson et al. first estimate the serial interval—the time between two cases in a transmission chain— and then use a causal filter to estimate *R*(*t*). Chowell et al. compare four methods for estimating *R*(*t*) of the Spanish flu in San Francisco from 1918 to 1919 [5].

The rapid spread of COVID-19 across the globe has led to tremendous research efforts on estimating reproduction numbers. For instance, the German RKI relies on nowcasting to estimate the value of *R*(*t*) [2]. It uses nowcasting to account for reporting delays, and it imputes missing values. Other papers focus on the effect of interventions. For instance, Khailaie et al. examine the impact of interventions on the reproduction number *R*(*t*) in Germany [12], Hyafil and Morina focus on Spain [10], and Leung et al. estimate *R*(*t*) in various Chinese cities and provinces [13]. Additionally, Leung et al. evaluate different intervention strategies. The results from [4] indicate that the changes in *R*(*t*) are caused directly by interventions instead of gradual changes in the behavior. Dehning et al. combine Bayesian frameworks and an SIR-model to evaluate the time dependence of the effective growth rate [7]. Additionally, Dehning et al. identify three change points in the growth rate of COVID-19 in Germany—all of which correspond to governmental interventions [7]. Zhang et al. provide a similar study that analyzes the effect of interventions, but the work is extend to examine the interplay between age, contact pattern, social distancing, risk of infection, and infectioness [19]; for their work the authors conducted surveys in Wuhan and Shanghai. Park et al. compute the basic reproduction number with uncertainties [15], and Tariq et al. estimate the reproduction number by analyzing clusters of cases as well as by case incidences [17].

There are other studies inspired by the problems of delay patterns and/or misreporting. Azmon et al. evaluate *R*(*t*)-estimators by using simple artificial delay patterns [3]. Another example is the work of Abbott et al., which aims to reduce the effect of delays. To do so the authors use a delay—extracted from historical data—to calculate the onset date out of the date of the report. However, the authors do not consider weekday-dependent periodicities [1].

It should be noted that there often exists the assumption that nowcasting approaches implicitly mitigate weekly delay patterns. However, this is only the case if the nowcasting is nearly perfect. Furthermore, often filters with a length of one week are used for coping with periodic behavior of the delays. However, in most cases, the impact on the delays for fast changes of the reproduction number *R*(*t*) is not examined.

## 3 Estimating *R*(*t*) via Filtering

Broadly speaking, there are two paradigms to computing reproduction numbers: employing “compartmental models” or “time-since-infection models” [9]. We follow the latter approach as described in [9], according to which there exists the following relation between the number of cases *N* (*t*) at time *t* and the (instantaneous) reproduction number *R*(*t*)

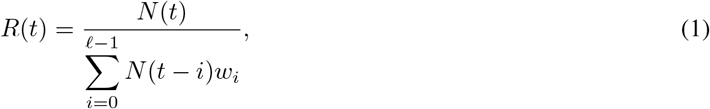

where *w*_*i*_ is the discretized distribution of the so-called generation time such that ∑_*i*_ *w*_*i*_ = 1, cf. [9]. The simplest estimate of the weights *w*_*i*_ is

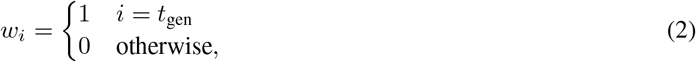

where *t*_gen_ is the so-called generation time [14, 11].

Estimators based on Equation (1) alone suffer from periodicities—which often occur in time-series. Thus, many approaches seek to reduce the influence of periodicities, for example, by using filters with the size of one typical period, e.g. a week.

In the following, we present one existing approach which uses causal filtering. Afterwards, we propose two further approaches which are based on acausal filtering. Figure 2 shows the connection between the three approaches: the causal filter is based on data from the past and the present; the acausal filter is based on data from the past, the present, and the future, but there are no means to determine the future values from the present; the projected filter uses data from the past, the present, and the future, and future values are projected from the past.

**Figure 2:**
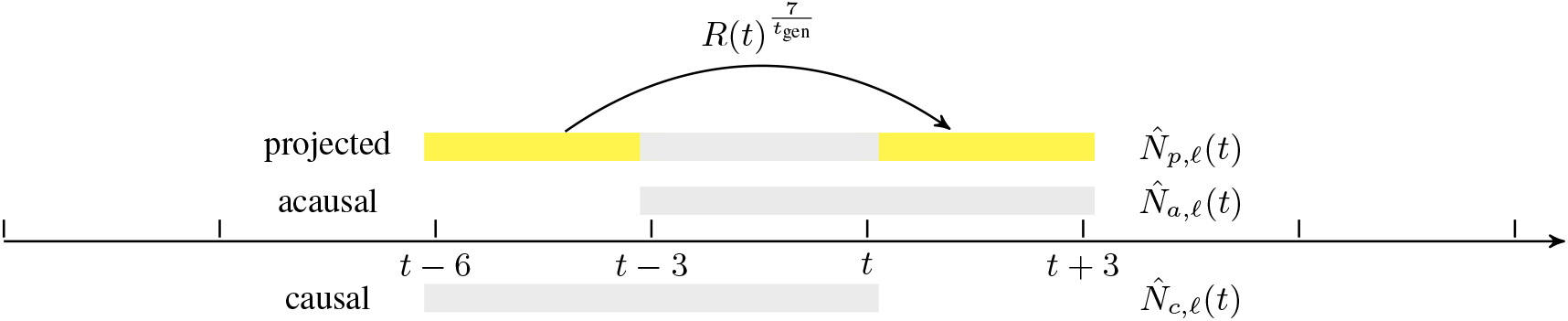
For the causal estimation of *R*(*t*) the cases of the past seven days are used. For the acausal estimation the cases of the past three days, the present day, and the three next days are used. To obtain the cases in the future we project the cases from the days *t* − 7 until *t* − 3 in the future, by using 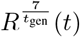. The filter length ℓ is 7.

All three approaches are inspired by Equation (1) and have the form

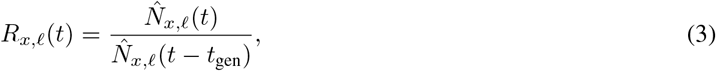

where *t* is the time, *t*_gen_ is the generation time, is the filter length, 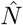 is the nowcasted number of cases, and the subscript *x* ∈ {*c, a, p*} determines the approach used for filtering.

- *c* specifies the causal filtered number of cases,
- *a* is the acausal filtered number of cases, and
- *p* is the projected and acausal filtered number of cases,

see also Figure 2. Note, since the true number of cases (*N*) is not known (compare Figure 1), the estimators have to work with the nowcasted number of cases 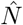.

From Equation (3), we see that the reproduction number is a quotient of case numbers. We compare three different filters of computing the number of cases.

The first filter for the total number of cases

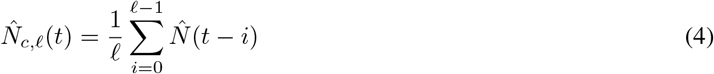

is the causal moving average of the estimated number of cases 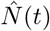 at time *t* over the last ℓ days. This method is used by [11]. Although this approach accounts for periodicities, it introduces unnecessary lag to the estimator (shown later).

A possible remedy is to employ an acausal filter, which uses not just the past and present but also future values for the number of cases. The acausal filtered 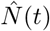^2^ is

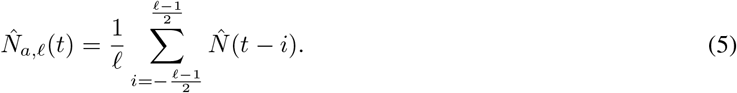

This is a symmetric filter; the day of interest is “in the center” of the filter.

The previous approach takes the future values for granted. Consequently, this approach is only suitable for a retrospective analysis of *R*(*t*). This method is used by [11]. To calculate the daily-based instantaneous reproduction number *R*(*t*) it remains to answer how to obtain future values. Therefore, we propose to project values from the past into the future. The resulting filtered number of cases is

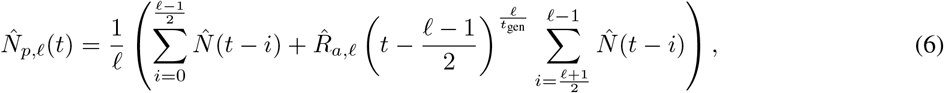

where 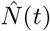 is the estimated number of cases at time *t*, 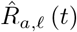, *ℓ* (*t*) is the acausally estimated reproduction number at time *t*, and *t*gen is the generation time. The term 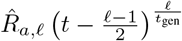 projects the value of the previous period into the next period. Note that it is possible to use the acausal variant 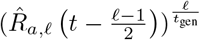, because at time *t* all values needed by 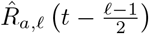 are available.

Having introduced three different filters for the number of cases, there follow three different estimators for the instantaneous reproduction number *R*(*t*)^3^.

*Remark* (Notation). In the following, we use either the Equation (3) either with the nowcasted number of cases 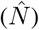 or the reported cases (*C*(*t*)). To facilitate distinction later on we use the superscripts *N* or *C* in combination with *R*(*t*). Also, we use *N* ∗ to denote the most current and consequently most inaccurate nowcasting method.

## 4 Creating Synthetic Datasets based on COVID-19 Data in Germany

What are meaningful ways to compare the three estimators from the previous Section 3? Clearly, applying them to real-world data is favorable. However, real-world data comes with real-world fuzziness. Hence, we suggest a method to generate synthetic data. Such approaches are popular in machine learning if the dataset is not good enough, e.g. Street and Kim created a synthetic dataset for getting a ground truth for concept drift detection [16].

For our purposes, the synthetic dataset needs to satisfy three requirements.

1. The ground truth shall be unambiguous. Hence, we create our dataset based on the given values of *R*(*t*).
2. The dataset shall contain realistic periodicities. Hence, we apply delay structures to the created cases that we extract from the publicly available COVID-19 data for Germany.
3. The dataset shall be realistic and it shall follow a curve similar to the reported cases.

The following algorithm generates a synthetic dataset fulfilling the above requirements; it is based on Equation (1). As input, the algorithm needs a smooth time-series of the reproduction number, the number of cases for the initial week, and a delay pattern. Based on this input, the algorithm performs the following two steps.

1. Based on previous values, generate new cases by rearranging Equation (1)

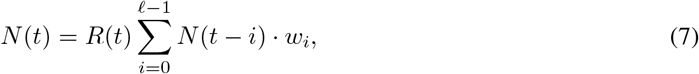

where *N* (*t*) is the number of new cases at time t, *w*_*i*_ weights how much infections are caused by cases at time *t* − *i*, and *R*(*t*) is the reproduction number at time *t*.
2. Afterward, apply a delay pattern to *N* (*t*) to obtain the cases with periodicities 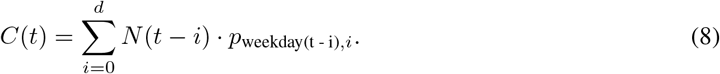

where *p*_weekday(t - i),*i*_ is the share of days with a delay of *i* at the weekday of time *t* + *i, t* is the time in days, and *d* is the longest considered delay. Note that *C*(*t*) is simplified since we do not consider further influences like testing rates, probabilities of false-negative (or false-positive) test results.

As mentioned above, the data generation needs a delay structure as input. Instead of using a one-day, two-day, or weekend delay structure as in [3], we extract them from real COVID-19 data since we assume that real-world delay structures ones are more complex. Therefore, a dataset containing the date of illness and the date of the report for cases is necessary. Given such data, the following steps extract a delay structure^4^.

1. For each combination of date of illness and record, calculate the difference between both.
2. For each date of illness, extract the weekday—we assume a weekly periodicity.
3. Group the data by the weekday and the delay. Afterwards, calculate the sum for each group. This sum is the absolute number of cases that fall ill on the particular weekday with a specific delay.
4. For each sum, divide it by the number of people who fall ill on the specific weekday. The result is the share of the cases for the particular weekday that is reported with the corresponding delay.

In the following, we describe the relevant parameters used for creating a dataset. As *R*(*t*) we choose a smoothed step sequence with 300 days. The three steps are one drop from 3.2 to 0.9 in step 30 and two increases from 0.9 to 0.95 at step 100 and from 0.95 to 1.0 at step 200. This step sequence is smoothed by

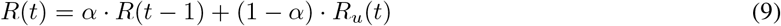

where *α* is a smoothing factor, *t* is the time, and *R*_*u*_(*t*) is the un-smoothed reproduction number.

The data generation also requires a delay structure. We extract it from the data provided by RKI5. This dataset contains cases of all German counties from January 28 until May 21. The following preprocessing steps are required^6^.

- Aggregate the data spatially for getting all cases of Germany for the specific date of illness and date of the report.
- Filter the data according to the following rules.
  – Consider only values for which the date of illness is reported.
  – Consider only values for which the date of illness is before the date of report.
  – Consider only values for which the difference between report date and date of illness is smaller than two weeks. The average delay in the complete dataset is 6.78 days. The average delay in the dataset with a maximum delay of two weeks is 5.48.

Figure 3 shows the resulting delay structure. Note that the sum for each weekday is one and that 10.16 % of the reported cases have a delay greater than two weeks. We observe that each weekday has a weekly delay structure. Moreover, the delay structure is different for each weekday. A probable explanation is the weekend’s strong influence on the periodicities. Also, the plot indicates that most cases are delayed for three or four days.

**Figure 3:**
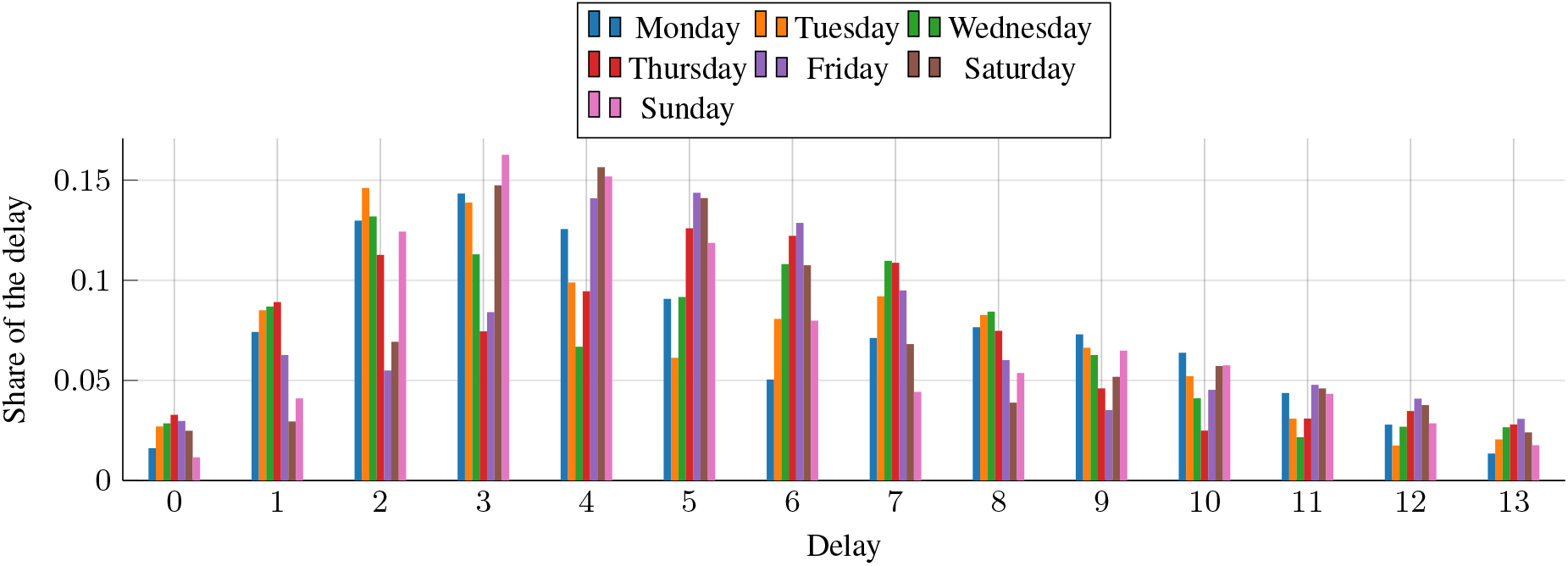
The delay pattern extracted from the COVID-19 data in Germany. The x-axis shows the delay in days and the y-axis the share of this delay for a specific weekday.

Finally, the last input needed by the data generation is the number of cases in the initial six days. For this parameter, we chose [1,2,2,3,3,4]. With these parameters we generate the synthetic dataset; it is shown in Figure 4. The analysis with a cross-correlation indicates that the curve of *C*(*t*) is delayed by ten days. Note that the delay between the peaks in the nowcasted numbers and the daily reports of the RKI is twelve days. The difference can be explained by additional delays that can arise from the forwarding of data by local health authorities to the central institution in Germany, the Robert Koch Institut (RKI).

**Figure 4:**
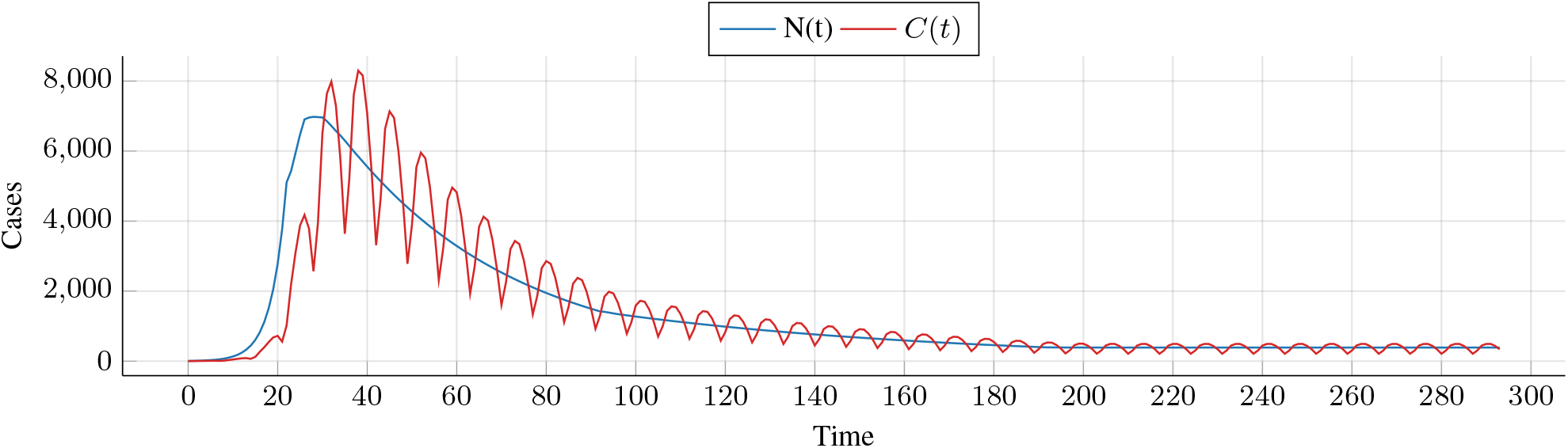
The number of cases and the reported cases of the artificially generated dataset are shown. The dataset is created with the extracted delay pattern (see Figure 3) and the *R*(*t*) of Equation (9)

## 5 Evaluation and Discussion – Synthetic data

In this subsection, we evaluate the estimators on the synthetic data generated in Section 5. Therefore, we test both proposed acausal estimators on the number of *N* (*t*) and the number of reported cases *C*(*t*), see Figure 1. The benchmark estimators are the ones from the RKI with filter lengths of four and seven [11].

### Accuracy of the *R*(*t*)-Estimators

We use the mean absolute error (MAE) to evaluate the accuracy of the three different estimators from Section 3

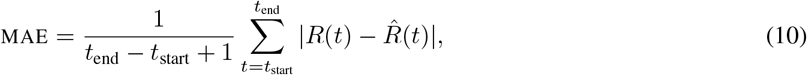

where *R*(*t*) is the reproduction number 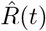 is the estimated reproduction number. The MAE is applied on the estimation of *R*(*t*) for the whole dataset, as well as on a truncated dataset without the first 55 values. The truncated dataset is used for evaluating the estimators without the starting phase, which dominates the MAE on the complete dataset.

Table 1 provides the MAE of the estimators on the synthetic dataset. The left column gives the accuracy on the reported data, the column in the middle on the reported data shifted by ten days, and the right column on *N* (*t*). Note that the delay of ten days is the delay between *C*(*t*) and *N* (*t*) measured by the cross-correlation.

**Table 1:** MAE of estimators on the full synthetic dataset, and MAE on the dataset without the starting phase; for reported cases, delayed reported cases, and real cases.

| On reported cases $C(t)$ | | | On reported cases $C(t)$ (delayed) | | | On real cases $N(t)$ | | |
| --- | --- | --- | --- | --- | --- | --- | --- | --- |
| Method | $t_{\text{start}} = 0$ | $t_{\text{start}} = 56$ | Method | $t_{\text{start}} = 0$ | $t_{\text{start}} = 56$ | Method | $t_{\text{start}} = 0$ | $t_{\text{start}} = 56$ |
| $\hat{R}_{c,4}^C(t)$ | 0.2891 | 0.2353 | $\hat{R}_{c,4}^C(t)$ | 0.2900 | 0.2359 | $\hat{R}_{c,4}^N(t)$ | 0.0154 | 0.0008 |
| $\hat{R}_{c,7}^C(t)$ | 0.0737 | 0.0078 | $\hat{R}_{c,7}^C(t)$ | 0.0550 | 0.0057 | $\hat{R}_{c,7}^N(t)$ | 0.0228 | 0.0015 |
| $\hat{R}_{a,7}^C(t)$ | 0.0410 | 0.0066 | $\hat{R}_{a,7}^C(t)$ | 0.0705 | 0.0062 | $\hat{R}_{a,7}^N(t)$ | 0.0085 | 0.0006 |
| $\hat{R}_{p,7}^C(t)$ | 0.0630 | 0.0055 | $\hat{R}_{p,7}^C(t)$ | 0.0452 | 0.0033 | $\hat{R}_{p,7}^N(t)$ | 0.0228 | 0.0013 |

### MAE for Reported Cases

*C*(*t*) For the reported cases on the complete data, 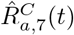 is the most accurate estimator. However, on the dataset without the starting phase, 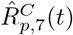 is more accurate. The reason, therefore, is that 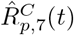 predicts the next value more accurately during stable periods. However, during periods where *R*(*t*) changes, it performs worse than 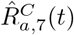. For the complete as well as the truncated dataset, the worst result is achieved by 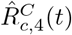.

### MAE for Delayed Reported Cases

*C*(*t*) For the delayed reported cases, the best result is achieved by 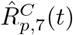, followed by 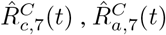, and 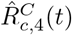. Moreover, we observe that the values of 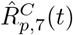 and 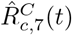 are improved compared to the non-delayed data. In contrast, the accuracy of 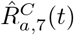 and 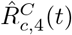 decreases. The reason, therefore, is that 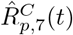 and 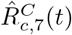 react slower than the other estimators. Consequently, the shift reduces the delay, and the accuracy improves.

### MAE for Real Cases

*N* (*t*) For *N* (*t*), the best model for MAE and truncated MAE is 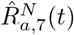, followed by 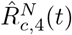. In contrast to the result on 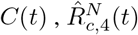 is better than 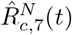 and 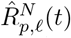. The reason, therefore, is that *N* (*t*) does not have weekly periodicities, which can harm accuracy. Consequently, the filter size does not influence the accuracy.

### Moving Average of MAE

For a more detailed examination of the accuracy, we evaluate the curves of the moving average of the MAE with a window of 14 days. Therefore, Figure 5 provides the complete curves as well as two cutouts. Note, we omit 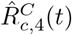 in the cutouts due to the high error. We make four observations.

**Figure 5:**
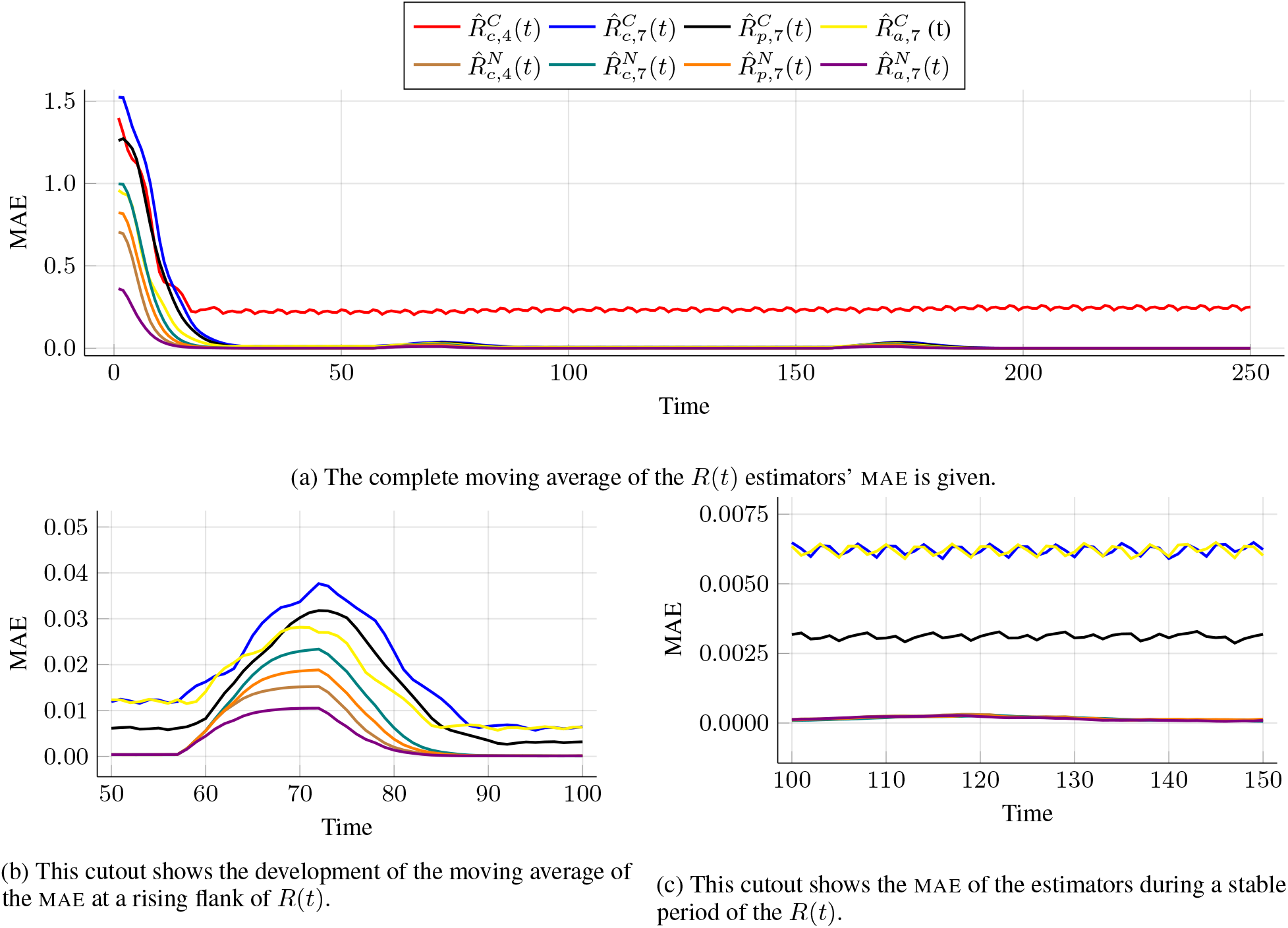
The moving average of the mean absolute error (MAE) of the different tested estimators is given. The window size for the moving average is 14. Note for both cutouts, we remove 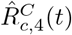 since its error is high, compared to the other estimators.

1. In Figure 5a, we observe that 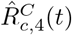 has a high error. In contrast, 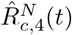 has a small error. The reason, therefore, is that 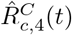 and 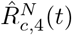 are prone to suffering from periodicities. However, *N* (*t*) has no periodicities in contrast to *C*(*t*). Consequently, only 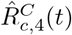 has a high error.
2. In Figure 5c, *R*(*t*) is stable, and so is the MAE of all estimators. However, the level of error differs.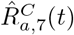 and 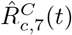 have higher error values than the 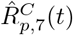. The estimators that bases on *N* (*t*) achieve the smallest.
3. During periods where *R*(*t*) changes (Figure 5b), all estimators have peaks in their moving MAE. The smallest error is achieved again by the estimators, which base on *N* (*t*). 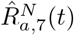 has the lowest peak, followed by 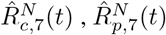, and 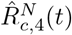. For the estimators on 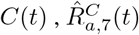 has the smallest peak. This observation indicates that 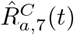 reacts faster than 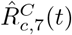 and 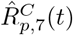.
4. Moreover, the MAE of all estimators based on *C*(*t*) is smaller after the change. This is caused by the fluctuations, which we analyze in the next part.

### Influence of Periodicities

Figure 6 shows the complete *R*(*t*) curves of the different estimators (Figure 6a) as well as two cutouts showing a rising flank of *R*(*t*) (Figure 6b) and a stable period (Figure 6c). We make five observations.

**Figure 6:**
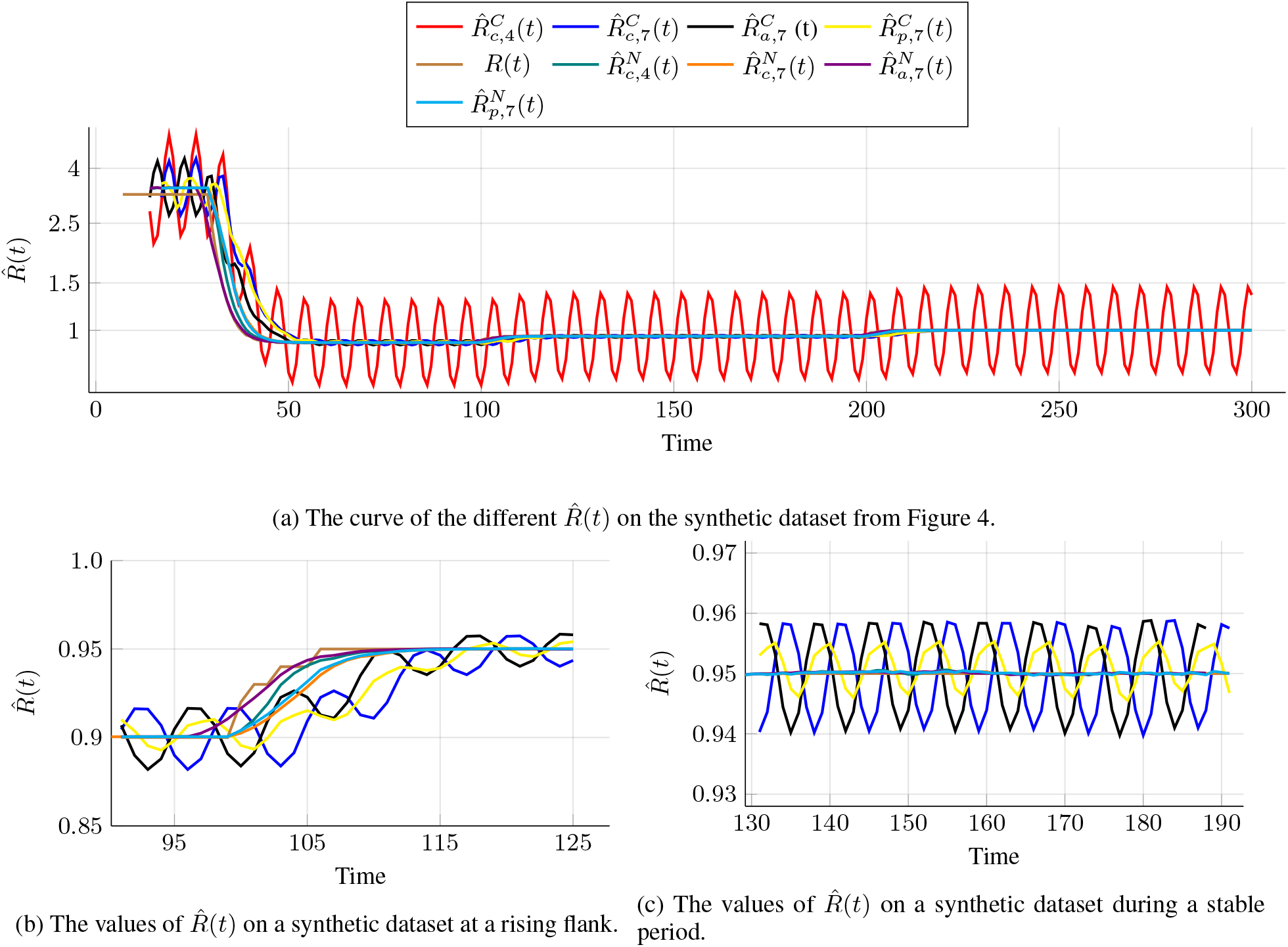
The 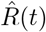 of different estimators and two cutouts are given. Note for both cutouts, we remove 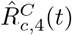 since it fluctuates stronger than the other estimators.

1. 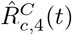 fluctuates stronger than the other estimators. The reason for this behavior is that this estimator works on data with weekly periodicities and that its filter size is not a multiple of one week. Thus, 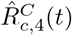 is not able to cope with the periodicities. Consequently, for making the cutouts clearer, we omit 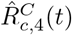 there.
2. Estimators that works on *C*(*t*) show different behavior than estimators on *N* (*t*). At the beginning of Figure 6 as well as during the complete cutouts (Figures 6b and 6c), we observe that the estimators that work on *N* (*t*) have fewer fluctuations than the estimators on *C*(*t*). The reason, therefore, is that *N* (*t*) does not have weekly periodicities. Accordingly, the corresponding estimators also have no fluctuations.
3. In Figure 6b, we observe that the estimators differ in their speed of capturing changes. Estimators based on acausal filtering adapt faster than estimators based on causal filtering. 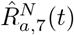 is even faster than the ground truth. The reason, therefore, is that acausal filter-based estimators consider future values.
4. In Figure 6b, we see that the fluctuations of estimators, which work on *C*(*t*) are smaller if the *R*(*t*) is nearer to one. This observation explains that if *R*(*t*) is one, the average number of new cases is constant. Consequently, if *R*(*t*) is one, variations of *C*(*t*) are only caused by weekly periodicities. On the other hand, an increasing distance of *R*(*t*) to one leads to a larger gap between the weekly minimum and weekly maximum of *C*(*t*). Thus, the estimated reproduction numbers fluctuate more if *R*(*t*) is further away from 1. This behavior also explains the remaining fluctuations, e.g., in Figure 6c.
5. The last observation is about the difference in the fluctuations of the estimators based on *C*(*t*). The amplitudes of the fluctuation of 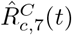 and 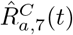 are the same. However, they differ in their phase. The cause of this behavior is the usage of causal and acausal filtering and the resulting shift. Moreover, we observe that 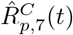 fluctuates less than 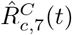 and 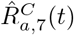. Probably, the cause of this behavior is that during phases of stable *R*(*t*) the 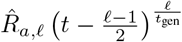 is countercyclical compared to 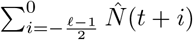, see Equation (6).

## 6 Use-case – COVID-19 Germany

In this section, we evaluate the performance of the estimators from Section 3 on a real dataset.

### COVID-19 Dataset for Germany

We use a daily updated dataset provided by the RKI7 for our use case. In the following, we use the version from the June 19, 2020. The first entry in the dataset pertains March 02, 2020; the last entry pertains June 18, 2020. This dataset contains different phases, e.g., an exponential increase of the number of cases (*R*(*t*) > 1) and a phase where R(t) fluctuates around one.

### Retrospective evaluation

Our first evaluation is on the retrospective evaluation of the course of COVID-19 in Germany. The tested methods and benchmarks are the same as in Section 5. Figure 7 shows the resulting curves of the estimated *R*(*t*). Note that the RKI has been using 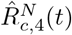 in its public communication since April 3, 2020. Since May 10, a new estimator was used in parallel based on 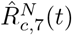, see [11]. We make four observations.

**Figure 7:**
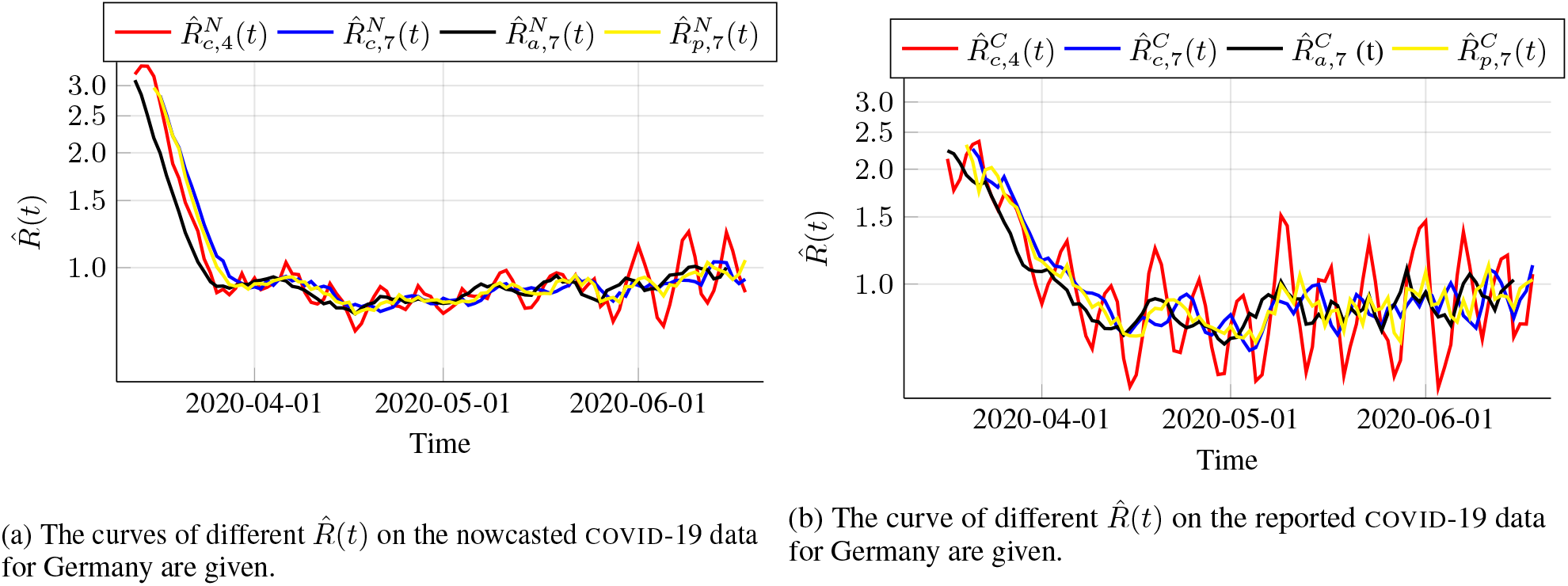
The curve of different 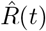 on the COVID-19 data for Germany are given.

1. The 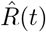 of all estimators decreases until the end of March 2020. Afterward, a period follows with estimates fluctuating around a stable constant follows. This is caused by the interventions against COVID-19 in Germany.
2. 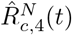 has stronger fluctuations than the other estimators. These fluctuations start to become even stronger at the beginning of June. Probably the reason is an accumulation of public holidays around the end of May and the beginning of June.
3. 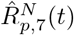 and 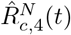 react faster to changes than the other estimators. We discussed this behavior already in the evaluation on the synthetic dataset (Section 5).
4. The last observation is that the estimators based on the reported data show higher fluctuations than the estimators on the nowcasted data. This observation is explainable by the nowcasting, which probably reduces the fluctuations, which are caused by the delay patterns.

### Evaluation of the Fluctuations

In the second evaluation, we investigate the fluctuations by comparing weekly minimal and maximal values of 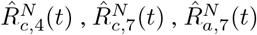, and 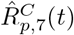 similar to [14]; the results are stated in Table 2. We make four observations.

**Table 2:**
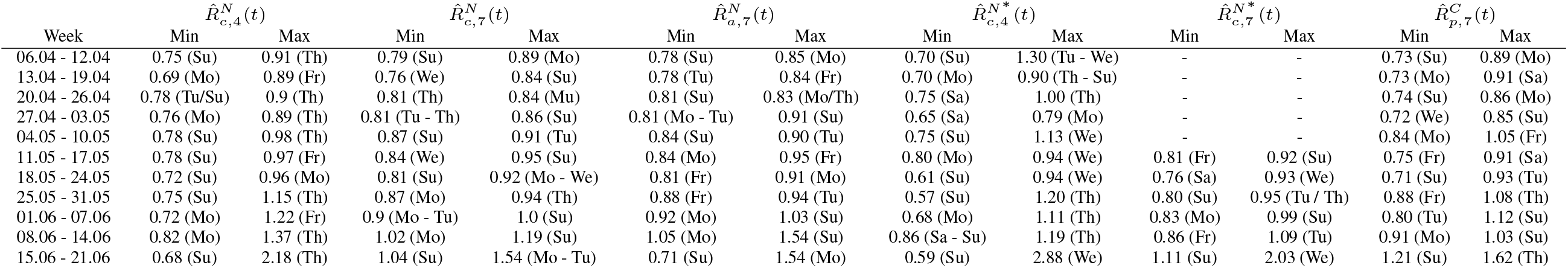
Minimal and maximal daily-based 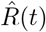 values for each week of a causal and an acausal *R*(*t*) estimator. The corresponding weekday is reported in the brackets. Note, the dates in this table refer to the date of illness, and the values of 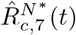 are only available from May, 10. Moreover, the asterisk denotes the most current and consequently most inaccurate nowcasting method.

1. We observe that the distance between minima and maxima 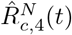 is larger compared to other estimators. Additionally, for 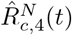, we see that the minima occur mostly on Sundays or Mondays. Such an accumulation is not observable for the other estimators. Probably, the higher fluctuations and the accumulation of the minima are caused by filtering over four days instead of filtering over a multiple of one week.
2. An additional observation is that the peak in 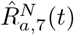 is before the peak in the other estimators. A possible cause is the acausal filtering on the nowcasted data. I.e., 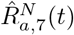 considers future values, which leads to a faster reaction to changes in *R*(*t*).
3. The estimators on the most recent nowcasting 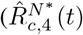 and 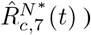 data fluctuate more than the estimators on the most accurate nowcasted data 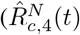 and 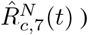. Probably, this behavior is caused by a smoothing effect of the nowcasting method used by the RKI.
4. 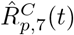 has a similar behavior as 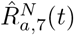 and 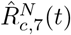. However, its fluctuations are slightly bigger. The reason is that 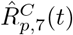 works on the *C*(*t*) data. Consequently, we can say that a retrospective acausal filtering on *C*(*t*) has a similar performance than causal and acausal filter-based *R*(*t*) estimators on nowcasted data. This observation indicates that 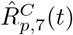 is an accurate *R*(*t*) estimator for the daily-based analysis.

## 7 Conclusion

We show that delays in reporting lead to fluctuations in *R*(*t*)-estimators. To overcoming this issue we propose to use filters with the length of one week. Moreover, we suggest to use acausal filtering instead of causal filtering, because this prevents the estimators from lagging behind the true reproduction number *R*(*t*). Acausal filters need future values. Hence, we project the values from the past into the future. Our results show that acausal filtering improves the overall accuracy and reduces fluctuations. Moreover, if the reproduction number *R*(*t*) is close to one, then the proposed approach is more robust and accurate than the benchmarks.

Another contribution of the present paper is to propose a methodology for synthetic data creation. It is undertaken on the basis of COVID-19 outbreak in Germany. The dataset contains all relevant real-world facets.

Future work will focus on improving synthetic data generation. For example, the data generation should consider under-reported cases (as in [3]), and herd immunity. Moreover, delays and the number of under-reported cases may change over time—aspects the data generation should consider. The longer COVID-19 continues to rage, the more societal aspects need to incorporated, e.g. public holidays and school vacation.

Also, with the diffusion ad smearing of COVID-19 across countries, the interpretability and relevance of *a single* reproduction number shrinks. Can *local* reproduction numbers (for counties or states) be devised, and how do they contribute to an improved *global* reproduction number?

## Data Availability

The data are provided by the Robert Koch Institue and are available at Github.

## List of Symbols

*C*(*t*): Number of reported/confirmed cases at time *t*
*t*gen: Generation time
*ℓ*: Filter length
*N* (*t*): Number of new cases at time *t*
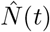: Estimated number of new cases (nowcasted) at time *t*
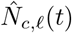: Causally filtered cases at time *t* for window length *L*
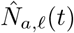: Acausally filtered cases at time *t* for window length *L*
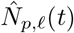: Acausally predicted and filtered cases at time *t* with window length *L*
*P*_weekday(t)_: Delay structure for the weekday of time *t*
*R*(*t*): Basic reproduction number at time *t*
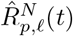: Acausal estimator for basic reproduction number at time *t* for window length *L*, based on the predicted number of cases
*w*_*i*_: Weights

## Acknowledgments

We would like to thank the RKI for providing both datasets used in the present paper.

## CRediT authorship Contribution Statement

**Benedikt Heidrich:** Data curation, Investigation, Software, Validation, Visualization, Writing - original draft. **Tillmann Mühlpfordt:** Data curation, Investigation, Software, Validation, Visualization, Writing -review & editing. **Veit Hagenmeyer:** Funding acquisition, Writing - review & editing. **Ralf Mikut:** Conceptualization, Data curation, Investigation, Methodology, Project administration, Validation, Writing - review & editing.

## Footnotes

1 Often, the current state of epidemic are not known or uncertain, e.g., the current number of cases is not immediately available. Therefore, these values are estimated based on past values. This is denoted as nowcasting.

2 For notational simplicity we assume that ℓ is odd.

3 An implementation of the estimators can be found at https://github.com/timueh/COVID-19

4 The code for the synthetic data generation is shared at https://github.com/timueh/COVID-19 in *synthetic-data/Synthetic_data*.*ipynb*

5 https://www.arcgis.com/home/item.html?id=f10774f1c63e40168479a1feb6c7ca74 version: May 22

6 The code for the delay pattern extraction is shared at https://github.com/timueh/COVID-19 in *synthetic-data/Extract true delay pattern*.*ipynb*

7 https://www.rki.de/DE/Content/InfAZ/N/Neuartiges_Coronavirus/Projekte_RKI/Nowcasting_Zahlen.xlsx?__blob=publicationFile

## References

[1] S. Abbott, J. Hellewell, R. N. Thompson, K. Sherratt, H. P. Gibbs, N. I. Bosse, J. D. Munday, S. Meakin, E. L. Doughty, J. Y. Chun, Y.-W. D. Chan, F. Finger, P. Campbell, A. Endo, C. A. B. Pearson, A. Gimma, T. Russell, S. Flasche, A. J. Kucharski, R. M. Eggo, and S. Funk. Estimating the time-varying reproduction number of SARS-CoV-2 using national and subnational case counts [version 1; peer review: awaiting peer review]. Wellcome Open Research, 5(112), 2020. doi: 10.12688/wellcomeopenres.16006.1. URL https://wellcomeopenresearch.org/articles/5-112/v1

[2] M. an der Heiden and O. Hamouda. Schätzung der aktuellen Entwicklung der SARS-CoV-2-Epidemie in Deutschland – Nowcasting. Epidemiologisches Bulletin, 2020(17):10–15, 2020. doi: 10.25646/6692.4. URL https://edoc.rki.de/handle/176904/6650.4.s

[3] A. Azmon, C. Faes, and N. Hens. On the estimation of the reproduction number based on misreported epidemic data. Statistics in Medicine, 33(7):1176–1192, 2014. ISSN 1097-0258. doi: 10.1002/sim.6015. URL https://onlinelibrary.wiley.com/doi/full/10.1002/sim.6015.

[4] M. V. Barbarossa, J. Fuhrmann, J. H. Meinke, S. Krieg, H. V. Varma, N. Castelletti, and T. Lippert. The impact of current and future control measures on the spread of COVID-19 in Germany. medRxiv, 2020. doi: 10.1101/2020.04.18.20069955. URL https://www.medrxiv.org/content/early/2020/04/24/2020.04.18.20069955.

[5] G. Chowell, H. Nishiura, and L. M. A. Bettencourt. Comparative estimation of the reproduction number for pandemic influenza from daily case notification data. Journal of the Royal Society, Interface, 4(12):155–166, 2007. doi: 10.1098/rsif.2006.0161. URL https://royalsocietypublishing.org/doi/10.1098/rsif.x2006.0161.

[6] A. Cintrón-Arias, C. Castillo-Chávez, L. M. A. Bettencourt, A. L. Lloyd, and H. T. Banks. The estimation of the effective reproductive number from disease outbreak data. Mathematical Biosciences and Engineering, 6(2): 261–282, 2009. doi: 10.3934/mbe.2009.6.261. URL https://www.aimspress.com/article/10.3934/mbe.2009.6.261.

[7] J. Dehning, J. Zierenberg, F. P. Spitzner, M. Wibral, J. P. Neto, M. Wilczek, and V. Priesemann. Inferring change points in the spread of COVID-19 reveals the effectiveness of interventions. Science, 2020. doi: 10.1126/science.abb9789. URL https://science.sciencemag.org/content/early/2020/05/14/science.abb9789.full.pd.

[8] S. Flaxman, S. Mishra, A. Gandy, H. J. T. Unwin, T. A. Mellan, H. Coupland, C. Whittaker, H. Zhu, T. Berah, J. W. Eaton, M. Monod, A. C. Ghani, C. A. Donnelly, S. M. Riley, M. A. C. Vollmer, N. M. Ferguson, L. C. Okell, and S. Bhatt. Estimating the effects of non-pharmaceutical interventions on COVID-19 in Europe. Nature, pages 1–8, 2020. doi: 10.1038/s41586-020-2405-7. URL https://www.nature.com/articles/s41586-020-2405-7.

[9] C. Fraser. Estimating individual and household reproduction numbers in an emerging epidemic. PLoS ONE, 2(8): 1–12, 2007. doi: 10.1371/journal.pone.0000758. URL https://journals.plos.org/plosone/article?id=10.1371/journal.pone.0000758.

[10] A. Hyafil and D. Morina. Analysis of the impact of lockdown on the evolution COVID-19 epidemics in Spain. medRxiv, 2020. doi: 10.1101/2020.04.18.20070862. URL https://www.medrxiv.org/content/10.1101/2020.04.18.20070862v1.

[11] R. K. Institut. Erläuterung der Schätzung der zeitlich variierenden Reproduktionszahl R, 2020. URL https://www.rki.de/DE/Content/InfAZ/N/Neuartiges_Coronavirus/Projekte_RKI/R-Wert-Erlaeuterung.pdf?__blob=publicationFile.

[12] S. Khailaie, T. Mitra, A. Bandyopadhyay, M. Schips, P. Mascheroni, P. Vanella, B. Lange, S. Binder, and M. Meyer-Hermann. Estimate of the development of the epidemic reproduction number Rt from coronavirus SARS-CoV-2 case data and implications for political measures based on prognostics. medRxiv, 2020. doi: 10.1101/2020.04.04.20053637. URL https://www.medrxiv.org/content/10.1101/2020.04.04.20053637v1.

[13] K. Leung, J. T. Wu, Di Liu, and G. M. Leung. First-wave COVID-19 transmissibility and severity in China outside Hubei after control measures, and second-wave scenario planning: a modelling impact assessment. The Lancet, 395(10233):1382–1393, 2020. doi: 10.1016/S0140-6736(20)30746-7. URL http://www.sciencedirect.com/science/article/pii/S0140673620307467.

[14] R. Mikut, T. Mühlpfordt, M. Reischl, and V. Hagenmeyer. Schätzung einer zeitabhängigen Reproduktionszahl R für Daten mit einer wöchentlichen Periodizität am Beispiel von SARS-CoV-2-Infektionen und COVID-19. URL https://dx.doi.org/10.13140/RG.2.2.14990.59207.

[15] S. W. Park, B. M. Bolker, D. Champredon, D. J. Earn, M. Li, J. S. Weitz, B. T. Grenfell, and J. Dushoff. Reconciling early-outbreak estimates of the basic reproductive number and its uncertainty: framework and applications to the novel coronavirus (SARS-CoV-2) outbreak. medRxiv, 2020. doi: 10.1101/2020.01.30.20019877. URL https://www.medrxiv.org/content/10.1101/2020.01.30.20019877v4.

[16] W. N. Street and Y. Kim. A streaming ensemble algorithm (SEA) for large-scale classification. In F. Provost, R. Srikant, M. Schkolnick, and D. Lee, editors, Proceedings of the Seventh ACM SIGKDD International Conference on Knowledge Discovery and Data Mining - KDD ‘01, New York, New York, USA, 2001. ACM Press. ISBN 158113391X. doi: 10.1145/502512.502568. URL https://dl.acm.org/doi/10.1145/502512.502568.

[17] A. Tariq, Y. Lee, K. Roosa, S. Blumberg, P. Yan, S. Ma, and G. Chowell. Real-time monitoring the transmission potential of COVID-19. medRxiv, 2020. doi: 10.1101/2020.02.21.20026435. URL https://www.medrxiv.org/content/10.1101/2020.02.21.20026435v7.

[18] R. N. Thompson, J. E. Stockwin, R. D. van Gaalen, J. A. Polonsky, Z. N. Kamvar, P. A. Demarsh, E. Dahlqwist, S. Li, E. Miguel, T. Jombart, J. Lessler, S. Cauchemez, and A. Cori. Improved inference of time-varying reproduction numbers during infectious disease outbreaks. Epidemics, 29:100356, 2019. doi: 10.1016/j.epidem.2019.100356. URL http://www.sciencedirect.com/science/article/pii/S1755436519300350.

[19] J. Zhang, M. Litvinova, Y. Liang, Y. Wang, W. Wang, S. Zhao, Q. Wu, S. Merler, C. Viboud, A. Vespignani, M. Ajelli, and H. Yu. Changes in contact patterns shape the dynamics of the COVID-19 outbreak in China. Science, (368):1481–1486, 2020. doi: 10.1126/science.abb8001. URL https://science.sciencemag.org/content/368/6498/1481.abstract.

